# A Mathematical prediction of the time evolution of the Covid-19 pandemic in some countries of the European Union using Monte Carlo simulations

**DOI:** 10.1101/2020.04.10.20061051

**Authors:** Ignazio Ciufolini, Antonio Paolozzi

## Abstract

In this paper we study the statistical evolution in time of the Covid-19 pandemic in Spain, Italy, Germany, Belgium, The Netherlands, Austria and Portugal, i.e., the countries of the European Union (EU) that have a number of positive cases higher than 12 thousand at April 7, 2020. France is the third country of the EU for number of cases but a jump in the data on April 3, 2020 does not allow, at least for the moment, to have a reliable prediction curve. The analysis is based on the use of a function of the type of a Gauss Error Function, with four parameters, as a Cumulative Distribution Function (CDF). A Monte Carlo analysis is used to estimate the uncertainty. The approach used in this paper is mathematical and statistical and thus does not explicitly consider a number of relevant issues, including number of nasopharyngeal swabs, mitigation measures, social distancing, virologic, epidemiological and models of contamination diffusion.

## 1. Introduction

In this paper the cumulative diagnosed positive cases of Covid-19 infections and fatalities in those countries of the European Union with a number of cases higher than 12 thousand are considered. The data have been collected in the web sites of the World Health Organization^1^, Worldometer^2^ and Italian “Ministero della Salute” ^3^. On the basis of our previous analyses of the cumulative diagnosed positive cases in China and Italy^4,5^, we found that the distributions of the positive cases and of fatalities can well be approximated by a Cumulative Distribution Function (CDF) of the type of the Gauss Error Function with four parameters. The Gauss Error Function is the integral of a normal, Gaussian distribution and has been used in similar studies^6,7,8^. It is assumed that the diagnosed cases are a good statistical representation of the entire population of the much larger positive cases. The number of fatalities is probably more reliable than the number of positive cases since they usually occur in the hospital. There is an obvious uncertainty also for those occurrences since there are fatalities, not occurring in the hospital, associated with several severe and/or chronic diseases in which coronavirus could have had a role but that could not be diagnosed.

We have analyzed the following seven countries of the European Union: Spain, Italy, Germany, Belgium, The Netherlands, Austria and Portugal. France, that is the third country for number of positive cases within the EU, however it was not reported here because the fitting of the data does not look too accurate due to a jump in the number of cases diagnosed around April 3, 2020. We have also performed 25000 of Monte Carlo simulations to evaluate the uncertainty in the evolution of the Covid-19 pandemic in those countries.

## 2. Evolution of cumulative diagnosed positive cases in some countries of the European Union

As done in the case of China and Italy^4,5^, we have fitted the cumulative numbers of positive cases with a function of the type of the Gauss Error Function:

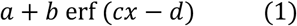

containing the four parameters a, b, c and d, to be fitted using the available official data for Spain, Germany, Belgium, The Netherlands, Austria and Portugal. Also, the fit of the Italian case is reported with an updated number of cases up to the April 7, 2020.

The results of these fits, that predict the evolution of the cumulative diagnosed positive cases in those countries, are reported in Figs. 1 to 4. In Fig. 1 and 2 the positive cases of Spain and Germany are compared with the ones in Italy. In Fig. 3 are reported the positive cases of Belgium and the Netherlands and in Fig. 4 the positive cases of Austria and Portugal.

**Fig. 1.**
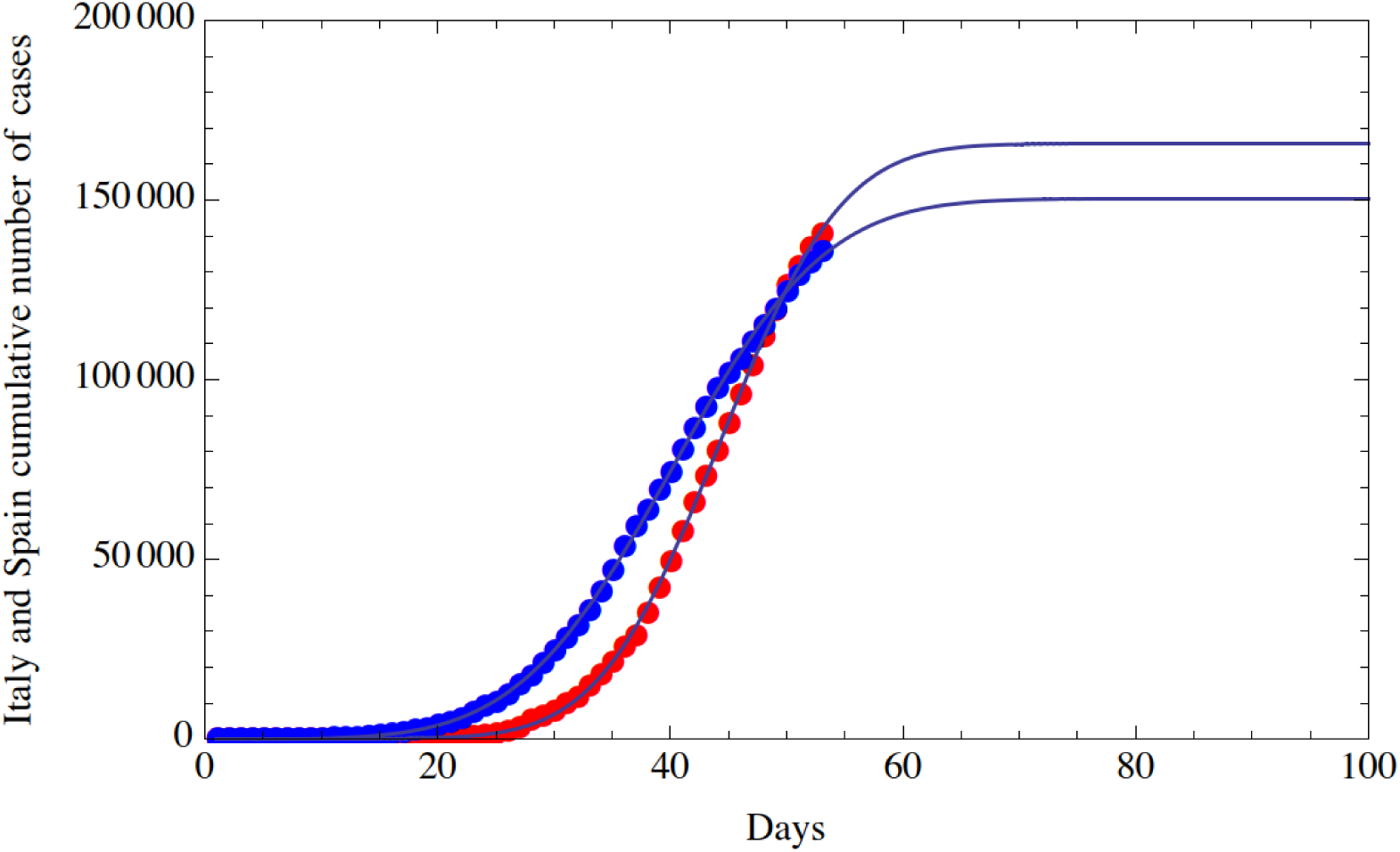
Fit of the cumulative number of diagnosed positive cases of Covid-19 in Spain (red dots) and in Italy (blue dots) from February 15, 2020 (included) to April 7, 2020 (included). The horizontal axis reports the days from February 15, 2020; the vertical axis reports the cumulative number of diagnosed positive cases.

**Fig. 2.**
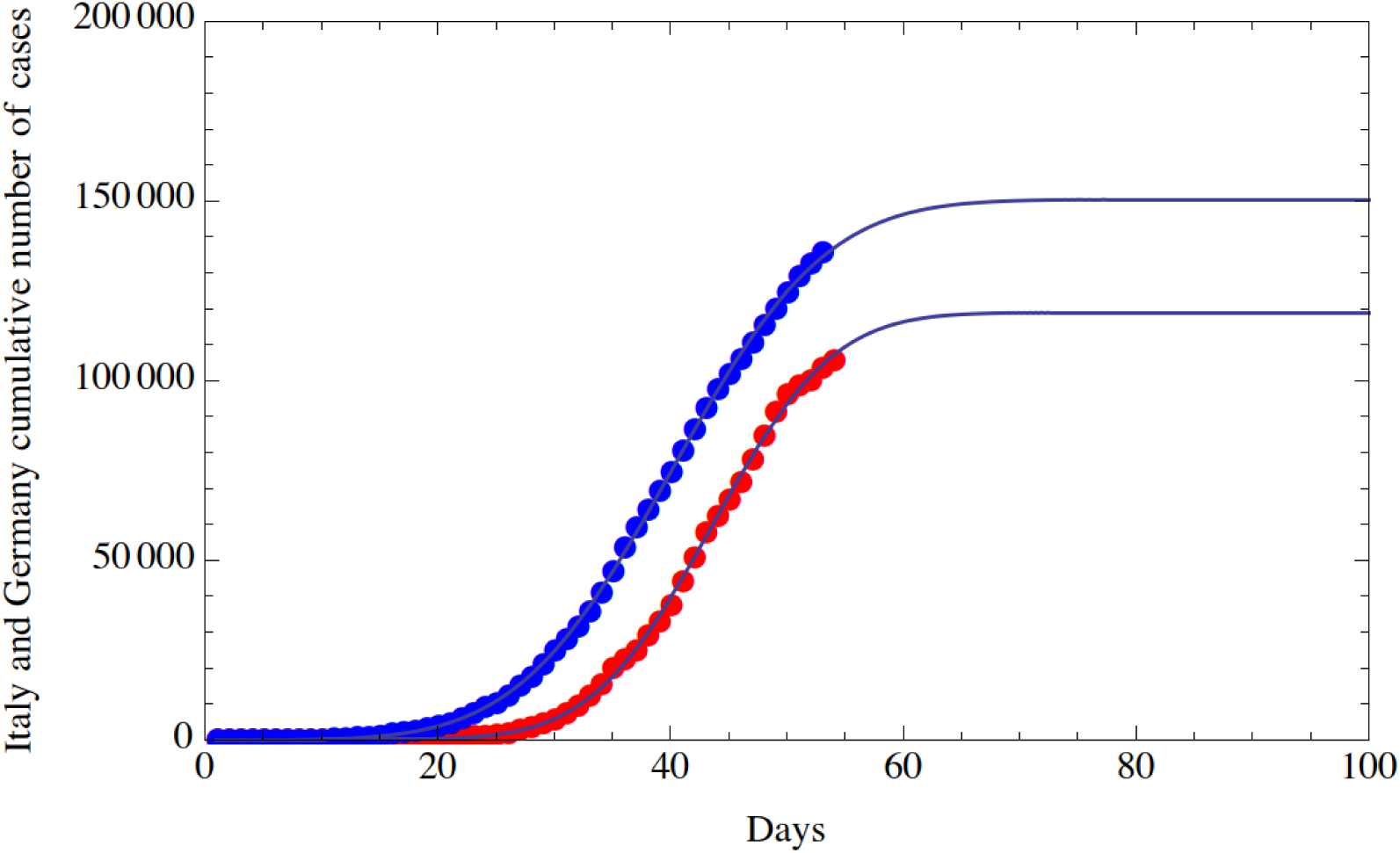
Fit of the cumulative number of diagnosed positive cases of Covid-19 in Germany (red dots) and in Italy (blue dots) from February 15, 2020 (included) to April 7, 2020 (included). The horizontal axis reports the days from February 15, 2020; the vertical axis reports the cumulative number of diagnosed positive cases.

**Fig. 3.**
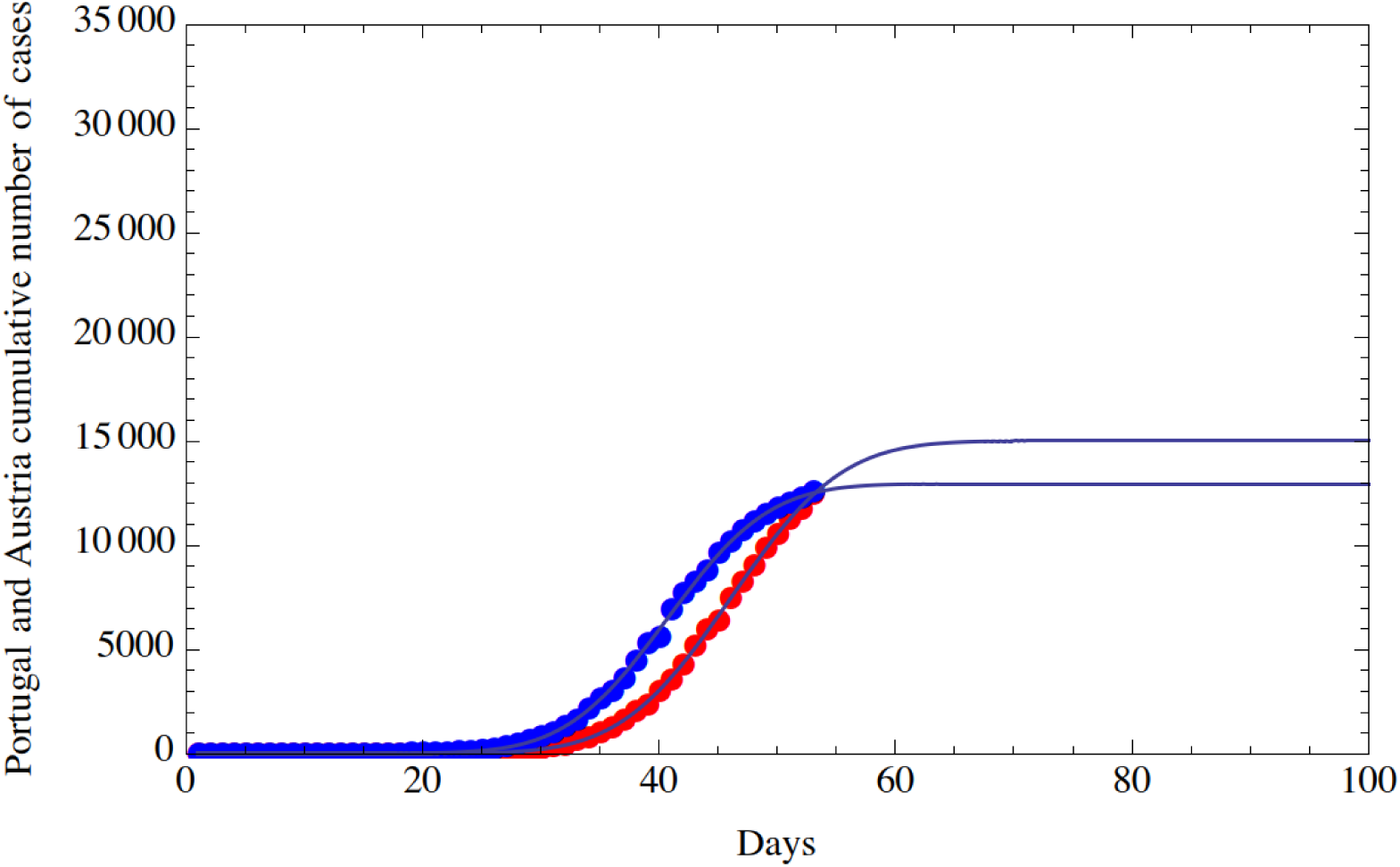
Fit of the cumulative number of diagnosed positive cases of Covid-19 in Belgium (red dots) and in the Netherlands (blue dots) from February 15, 2020 (included) to April 7, 2020 (included). The horizontal axis reports the days from February 15, 2020; the vertical axis reports the cumulative number of diagnosed positive cases.

**Fig. 4.**
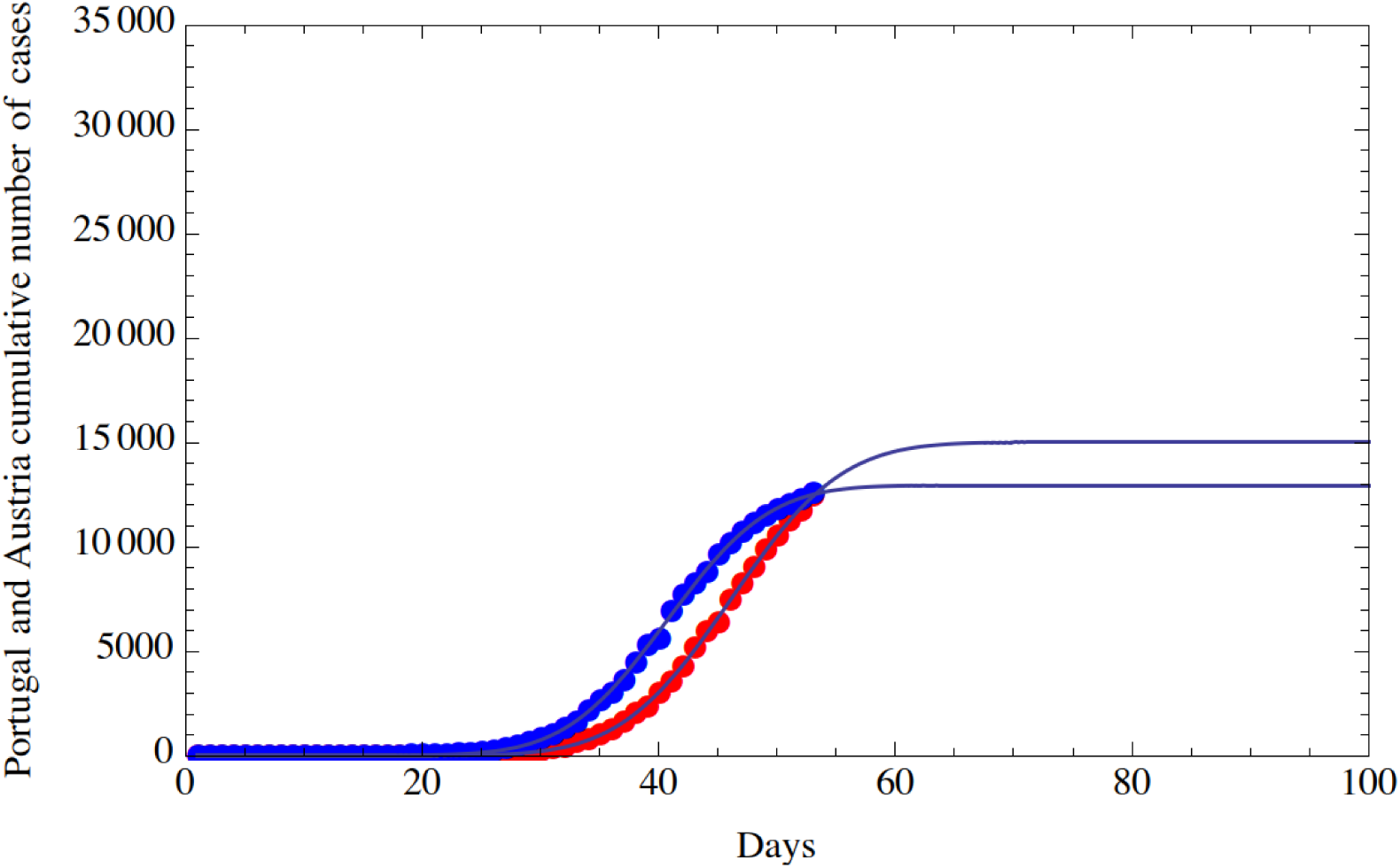
Fit of the cumulative number of diagnosed positive cases of Covid-19 in Portugal (red dots) and in Austria (blue dots) from February 15, 2020 (included) to April 7, 2020 (included). The horizontal axis reports the days from February 15, 2020; the vertical axis reports the cumulative number of diagnosed positive cases.

In Table 1 are reported the values of the flex and of the day in which a substantial reduction of the number of cases and fatalities are expected. The threshold is the value of positive cases or fatalities chosen to express a substantial reduction of Covid-19 pandemic. The flex expresses the day in which the daily positive cases or the daily fatalities reduce, it is sometimes referred to as the peak of the epidemic since after that day the daily occurrences decelerate. The values of the relevant standard deviations are calculated in the paragraph dedicated to the Monte Carlo simulations.

**Table 1:** Mathematical predictions (based on a function of the type of a Gauss Error Function, with four parameters) for the flex and for a substantial reduction of positive cases and fatalities for seven countries of the European Union. The day of a substantial reduction of cases to about the threshold values reported in columns 2 and 5 is reported in columns 4 and 7, respectively.

|  | POSITIVE CASES |  |  | FATALITIES |  |  |
| --- | --- | --- | --- | --- | --- | --- |
|  | Threshold | Flex | Substantial reduction | Threshold | Flex | Substantial reduction |
|  | (Positive cases per day) | (day) | (day) | (Positive cases per day) | (day) | (day) |
| **Spain** | 100 | 44 | 69 | 10 | 45 | 67 |
| **Italy** | 100 | 40 | 70 | 13 | 43 | 72 |
| **Germany** | 75 | 44 | 68 | 2 | 49 | 70 |
| **Belgium** | 16 | 48 | 74 | 2 | 57 | 84 |
| **The Netherlands** | 14 | 46 | 73 | 2 | 49 | 71 |
| **Austria** | 9 | 41 | 61 | 1 | N.A. | 62 |
| **Portugal** | 9 | 46 | 68 | 1 | N.A. | 65 |

The number of nasopharyngeal swabs-to-active cases ratio is in Italy 755445/94067=8.03 at April 7, 2020; if we consider the total of active, dead and healed (which is equivalent to the total number of people that is or has been infected), the number (at April 7, 2020) is 135586 providing a value of 755445/135586=5.57. So, we can reasonably assume that on average every 5.57 tests (nasopharyngeal swabs) 1 positive case is diagnosed. On April 7, the new positive cases were 3039. We have assumed (see section on the Monte Carlo simulation) an uncertainty in the daily data of 10% which we may consider half due to the uncertainty in the number of tests (nasopharyngeal swabs) that will be performed in the days to come and the other 5% due to other uncertainties. Therefore, it may be acceptable in the case of Italy a variation in the number of daily tests of less than 0,05*3039*5,57=846 tests/day. A higher rate in the daily variation of tests will introduce a systematic bias that could bring the prediction outside the 2-sigma or even 3-sigma range. The systematic bias becomes more severe if all the changes are monotonic (as it seems the case in Italy where the number of daily nasopharyngeal swabs is increasing). Analogous calculation can be done for the other countries that we do not report here for brevity. Consequently, the numbers reported in the table as +/- 2-sigma or +/- 3-sigma are valid with the limitations just described. In simpler words, the predictions reported assume, among other things, that the number of tests (nasopharyngeal swabs) does not change too much during the evolution of the pandemic and does not constantly increase or decrease. The number of fatalities is perhaps less affected by this parameter and maybe used as well to analyze the evolution of the pandemic.

## 3. Mathematical evolution of cumulative fatalities in some countries of the European Union

In this section we repeat the analysis using Eq. (1) for the cumulative fatalities attributed to Covid-19 that occurred in Spain, Germany, Belgium, The Netherlands, Austria and Portugal. Furthermore, the fit of the Italian data is updated to the April 7, 2020 with respect to what already published^4,5^.

The results of these fits, that predict the fatalities in those countries, are reported in Figs. 5 to 8. In Fig. 5 and 6 the fatalities of Spain and Germany are compared with the ones of Italy. In Fig. 7 are reported the fatalities of Belgium and the Netherlands and in Fig. 8 are reported the fatalities of Austria and Portugal.

**Fig. 5.**
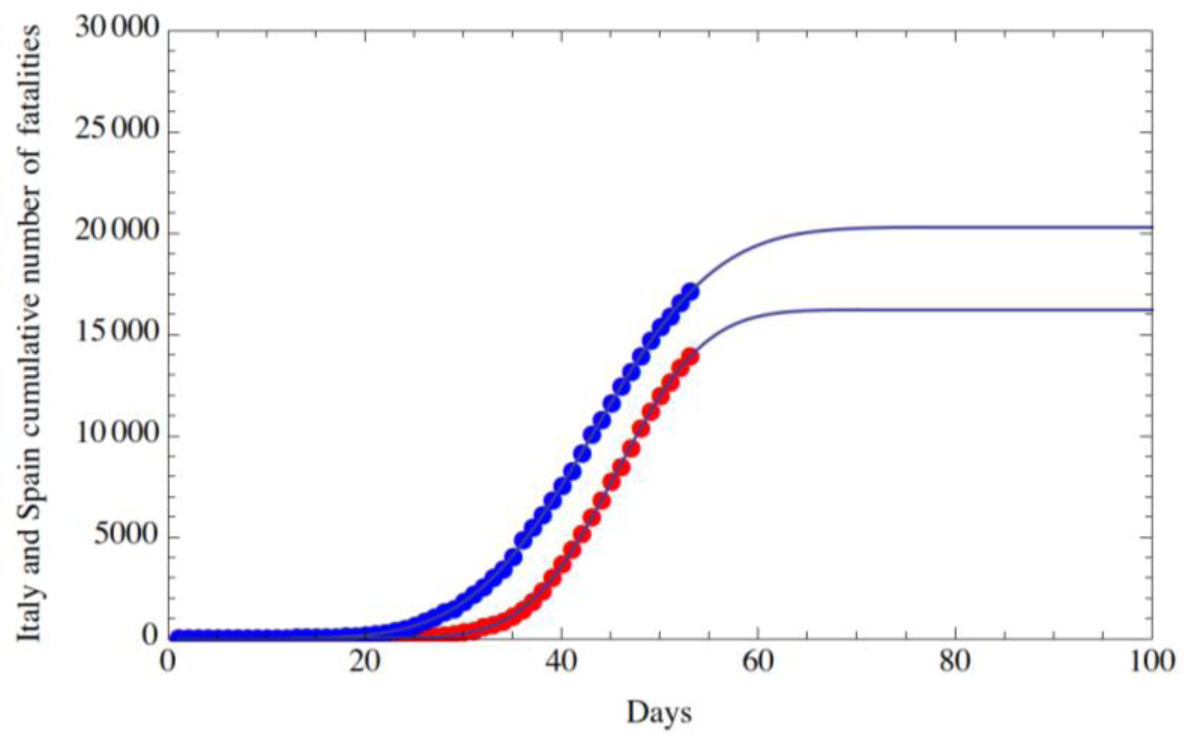
Fit of the cumulative number fatalities of Covid-19 in Spain (red dots) and in Italy (blue dots) from February 15, 2020 (included) to April 7, 2020 (included). The horizontal axis reports the days from February 15, 2020; the vertical axis reports the cumulative number of fatalities.

**Fig. 6.**
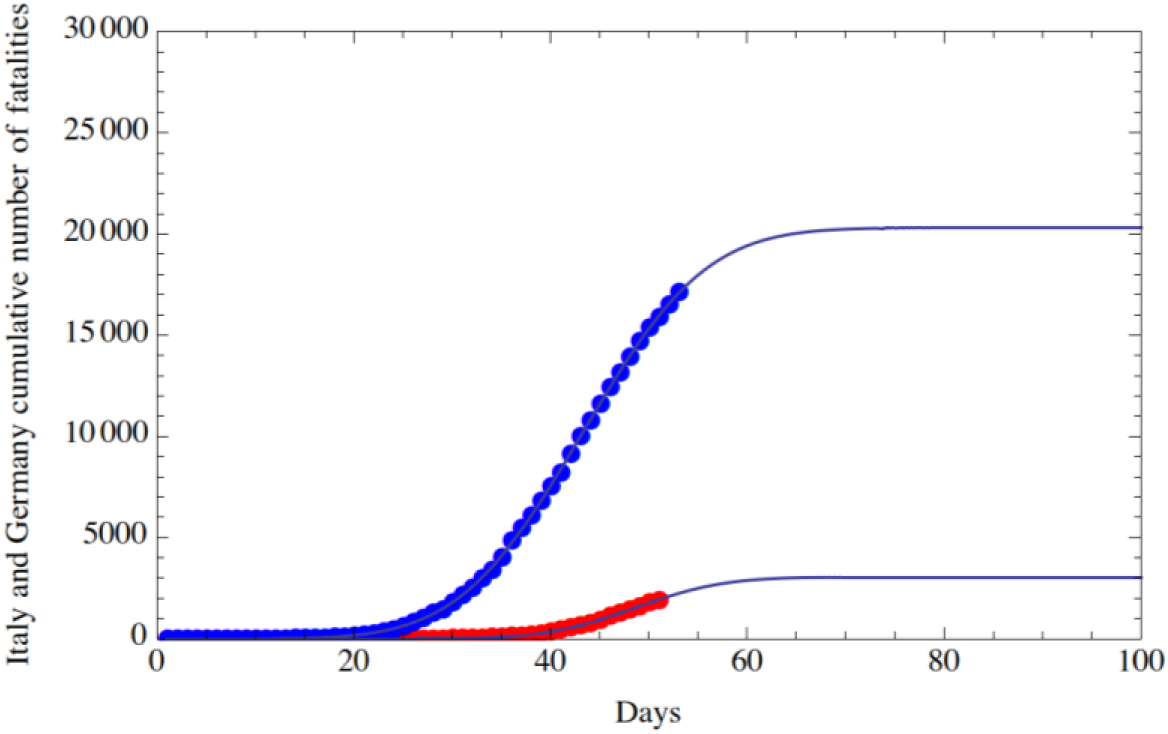
Fit of the cumulative number of fatalities of Covid-19 in Germany (red dots) and in Italy (blue dots) from February 15, 2020 (included) to April 7, 2020 (included). The horizontal axis reports the days from February 15, 2020; the vertical axis reports the cumulative number of fatalities.

**Fig. 7.**
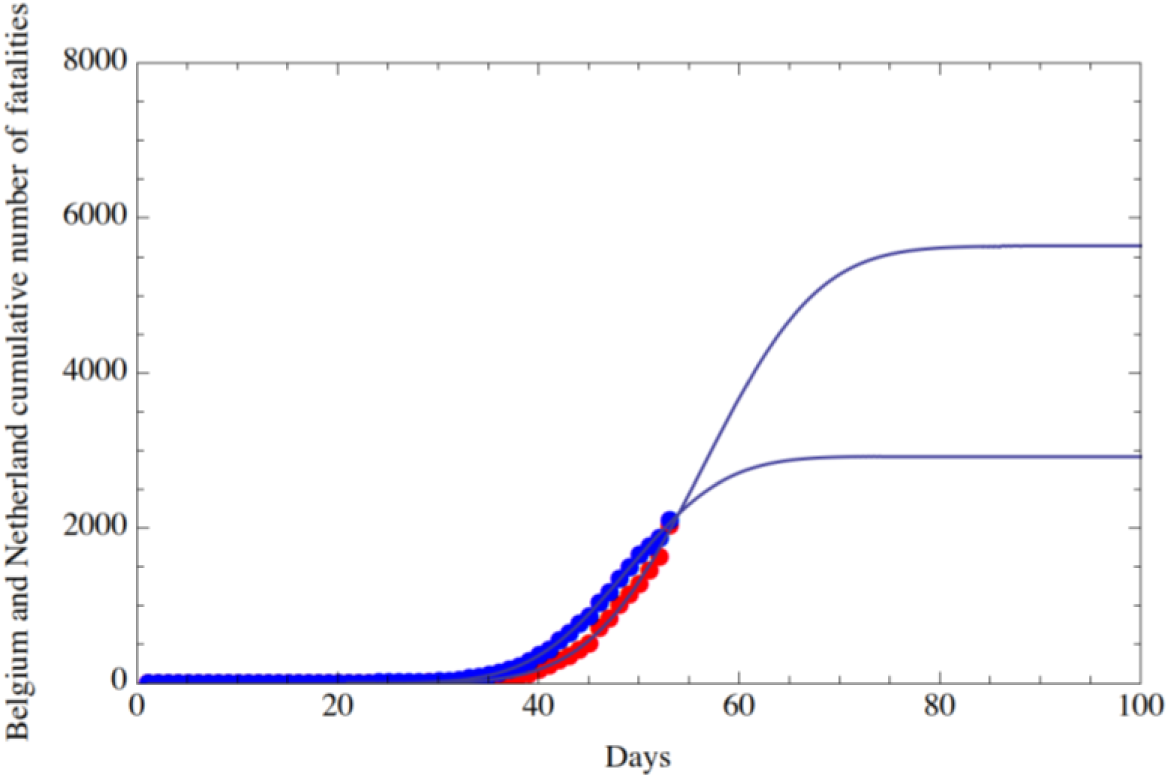
Fit of the cumulative number of fatalities of Covid-19 in Belgium (red dots) and in the Netherlands (blue dots) from February 15, 2020 (included) to April 7, 2020 (included). The horizontal axis reports the days from February 15, 2020; the vertical axis reports the cumulative number of fatalities.

**Fig. 8.**
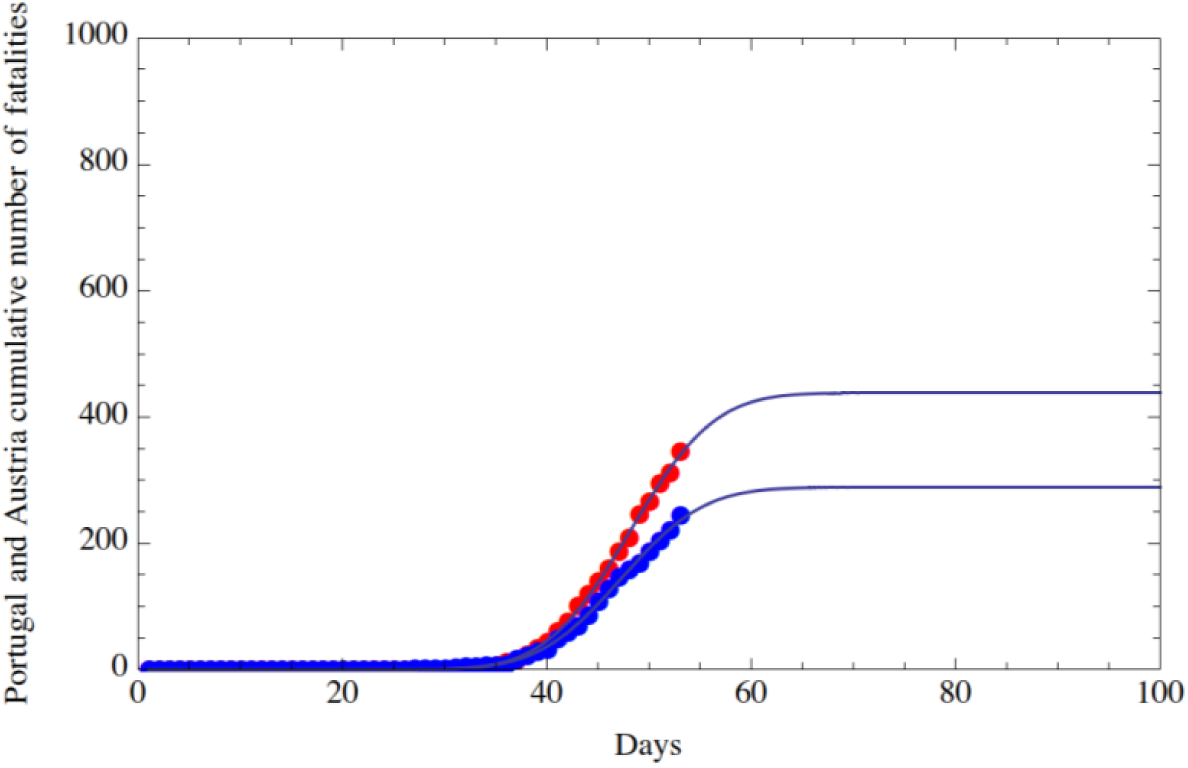
Fit of the cumulative number of fatalities of Covid-19 in Portugal (red dots) and in Austria (blue dots) from February 15, 2020 (included) to April 7, 2020 (included). The horizontal axis reports the days from February 15, 2020; the vertical axis reports the cumulative number of fatalities.

The crossing of the curves in Fig. 1 shows that Italy had a higher number of positive cases from the beginning. However, the number of positive cases in Spain started to grow faster as shown by the larger derivative of the red curve. This fast grow of cases in Spain was reported for several days in the news. Fig. 6 shows a much smaller number of fatalities for Germany and for this reason a substantial reduction of fatalities will occur a little earlier than for Italy. For all countries the flex point of the positive cases occurs always after Italy as one would expect. Austria is the country with the lowest number of cases (Fig. 8). In the case of the fatalities of Austria and Portugal, the numerical procedure did not converge so that the flex point is not available. Concerning the substantial reduction of positive cases, the threshold for the other EU countries, with respect to the value 100 chosen for Italy, was multiplied for a normalization factor taking into account the cumulative number of positive cases and of fatalities in Italy and in the other EU nations.

## 4. Monte Carlo simulations of cumulative positive cases and fatalities of Covid-19 in Italy

Several uncertainties can influence the diagnosed cases of the Covid-19 pandemic and in particular, the number of the daily positive cases is influenced by the number of nasopharyngeal swabs that can increase with time along with the awareness of the risks of the pandemic. For instance, Italy, in the days from April 4 to April 7, has reported an average of 33899 nasopharyngeal swabs while, on April 8, the number has increased by 51680.

To estimate the uncertainties in the number of positive cases and fatalities, we have used Monte Carlo simulations^9,10,11^ similarly to what done in previous works^4,5^. The uncertainty we consider in the Monte Carlo simulation is <u>not</u> the difference between the total number of the actual positive cases (which is unknown) and the diagnosed ones which can be one order of magnitude higher, or even more, than the actual cases. However, it is usual in statistics to use a sample as being representative of the population under study. The Monte Carlo simulations were performed both for the number of positive cases and the number of fatalities. For convenience to the reader we summarize here the procedure used previously^4,5^, the only difference being the number of simulations that have been increased from 150 to 25000. We have assumed a measurement uncertainty in the total number of positive cases and of fatalities equal to 10% of each daily number (Gaussian distributed).

Then, a random matrix (*m* × *n)* is generated, where *n* (columns) is the number of observed days and *m* (rows) is the number of random outcomes, which we have chosen to be 25000. Each number in the matrix is part of a Gaussian distribution with mean equal to 1 and sigma equal to 0.1 (i.e., 10% of 1), either row-wise and column-wise. So, starting from the *n* nominal values of the daily data, we generated *n* Gaussian distributions with 25000 outcomes, with means equal to the *n* nominal values and with 10% standard deviation. Then, for each of the 25000 simulations, those *n* values (corresponding to the cumulative positive cases, or fatalities, of *n* days) were fitted with a four parameter function of the type of the Gauss Error Function and we then determined the date of the flex with such fitted function for each simulation. Using the fitted function we also determined the date at which the number of daily positive cases will be less than a certain threshold that, for example, we have chosen for Italy and Spain to be 100 for the positive cases and 13 for the fatalities. Finally, we calculated the standard deviation of the 25000 simulations. In what follows, for the sake of brevity, we report only the figures relevant to the countries with the highest number of cases (Spain) and fatalities (Italy). The value of the standard deviation is about one day for all the simulations performed until the date of April 7.

In Fig. 9, in the case of Spain, is reported the histogram of the frequencies versus the day of a substantial reduction in the number of daily positive cases which has been chosen to be 100. The histogram approaches a gaussian with mean equal to day ≅ 69 and sigma of 1 day. This value of the mean is the same as obtained using the nominal values of the number of positive cases. In Fig. 10 is reported analogous results for the fatalities occurred in Italy up to April 7, 2020 (the threshold was chosen to be 13 of occurrences per day). Also, in this case the result is the same as obtained using the nominal values of the number of fatalities, i.e., 72.

**Fig. 9.**
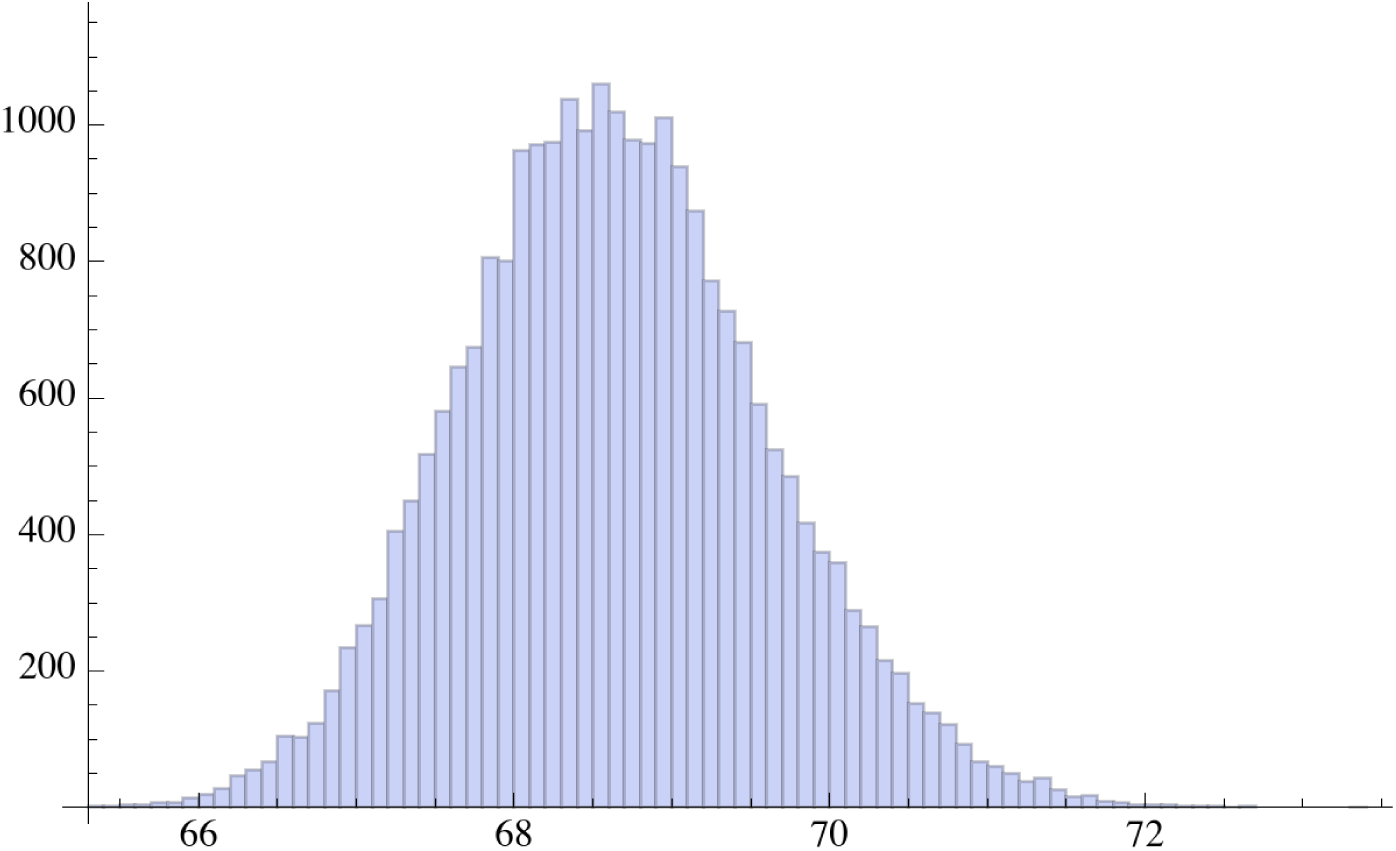
Monte Carlo simulations for Spain: histogram of frequencies versus day in which a substantial reduction in the number of daily cases (about 100) occur. The day number starts from February 15, 2020. The total number of simulations is 25000.

**Fig. 10.**
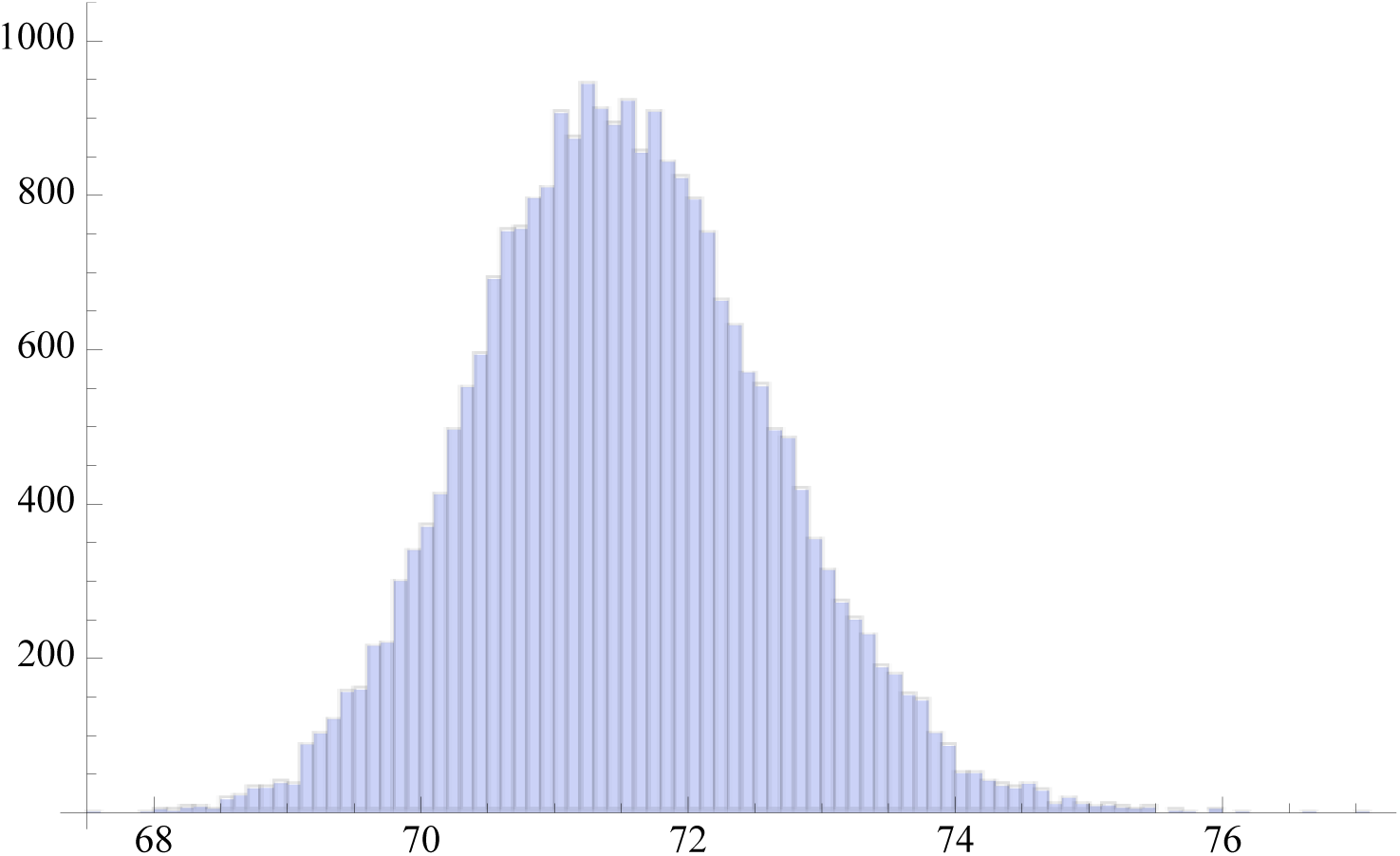
Monte Carlo simulations for Italy: histogram of frequencies versus day in which a substantial reduction in the number of fatalities (about 13) occur. The day number starts from February 15, 2020. The total number of simulations is 25000.

## 5. Conclusions

The mathematical predictions of the evolution of the Covid-19 pandemic in seven European Union countries are presented. The number of positive cases and of fatalities are fitted for 53 days from February 15, 2020 to April 7 2020. Several Monte Carlo simulations, each with 25000 runs, have provided an estimate of the standard deviation of the day of the flex and of the day in which a substantial reduction of positive cases and fatalities will occur. The function used for the fitting is of the type of the Gauss Error Function with four parameters. We obtained that the date of the flex, for all the countries considered here, occurs after Italy both for the positive cases and for the fatalities. The day of a substantial reduction in the number of daily positive cases or fatalities (which, for example, we took for Italy to be less than 100, while for the other countries such value was suitably scaled), does not always occur first for Italy. The predictions discussed in this paper are statistical in nature and do not explicitly take into account the relevant factors of number of nasopharyngeal swabs, social distancing, and epidemiological and virology studies, which are outside the analysis of the present paper.

## Data Availability

The data used for the paper are from publicily avilable repository and the sourced are listed in the references

## 6. Ackowledgements

We gratefully thank Richard Matzner (University of Texas at Austin) for helpful suggestions, Alessandro Paolozzi, and Claudio Paris (Centro Fermi).

